# Aerosol and droplet generation in upper and lower gastrointestinal endoscopy: whole procedure and event-based analysis

**DOI:** 10.1101/2021.04.15.21255544

**Authors:** Frank Phillips, Jane Crowley, Samantha Warburton, George S.D. Gordon, Adolfo Parra-Blanco

## Abstract

**Background and Aims:** Aerosol generating procedures have become an important healthcare issue due to the COVID-19 pandemic, as the SARS-CoV-2 virus can be transmitted via aerosols. We aimed to characterise aerosol and droplet generation in gastrointestinal endoscopy, where there is little evidence.

**Methods:** This prospective observational study included patients undergoing routine per-oral gastroscopy (POG, n=36), trans-nasal endoscopy (TNE, n=11) and lower gastrointestinal (LGI) endoscopy (n=48). Particle counters took measurements near the appropriate orifice (two models used, diameter ranges 0.3μm-25μm and 20μm-3000μm). Quantitative analysis was performed by recording specific events and subtracting the background particles.

**Results:** POG produced 1.96x the level of background particles (p<0.001) and TNE produced 2.00x (p<0.001) but a direct comparison shows POG produces 2.00x more particles than TNE. LGI procedures produce significant particle counts (p<0.001) with 2.4x greater production per procedure than POG but only 0.63x production per minute. Events significant relative to the room background particle count were: POG-throat spray (150.0x, p<0.001), oesophageal extubation (37.5x, p<0.001), coughing/gagging (25.8x, p<0.01); TNE-nasal spray (40.1x, p<0.001), nasal extubation (32.0x, p<0.01), coughing/gagging (20.0, p<0.01); LGI-rectal intubation (9.9x, *p*<0.05), rectal extubation (27.2x, *p* <0.01), application of abdominal pressure (9.6x, *p* <0.05), rectal insufflation/retroflexion (7.7x, *p* <0.01). These all produced particle counts larger than or comparable to volitional cough.

**Conclusion:** Gastrointestinal endoscopy performed via the mouth, nose or rectum all generates significant quantities of aerosols and droplets. As the infectivity of procedures is not established, we therefore suggest adequate PPE is used for all GI endoscopy where there is a high population prevalence of COVID-19. Avoiding throat and nasal spray would significantly reduce particles generated from UGI procedures.

## INTRODUCTION

Severe acute respiratory syndrome coronavirus 2 (SARS-CoV-2), the cause of the ongoing coronavirus 2019 (COVID-19) pandemic, can be transmitted via aerosols.^1-3^ Aerosol generating procedures (AGPs) therefore represent a transmission risk to healthcare workers and have become an important healthcare issue.^4^ Aerosols are in the respirable range, meaning they can deposit in the lower airways to cause infection via airborne transmission.^4^ In contrast, droplets are larger and gravitationally settle rapidly or can be inhaled at close contact, although resuspension into the air can occur from droplets on clothing or surfaces.^1^ The World Health Organization (WHO) have defined aerosols as particles <5μm and droplets as ∼5-10μm.^5^

The definition of an AGP lacks consensus: the WHO defines this as any medical procedure that can induce the production of aerosols of various sizes, including particles <5μm.^5^ However, Public Health England only considers AGP those resulting in release of airborne particles from the respiratory tract.^7^ Further difficulty with definitions arises since heavy breathing, talking, coughing, and singing all generate particles of varying sizes including aerosols.^8-9^

The WHO has produced a list of AGPs mainly based on evidence from small retrospective epidemiological studies linking these procedures with greater risk for healthcare worker infections.^10^ Aerosol or droplet levels were not measured in these studies, so the exact mode of transmission was not known. Although GI endoscopy is not on this list, various professional societies have designated upper GI endoscopy as an AGP, and LGI endoscopy at least of uncertain risk status, based on theoretical grounds.^11-14^ This has had important repercussions, including postponed procedures, lost capacity, and the use of enhanced PPE.

Two recent studies have provided evidence of aerosol generation during upper gastrointestinal (UGI) endoscopy using handheld particle counters. Chan *et al* showed that aerosols are generated during UGI endoscopy, and that continuous suction reduced aerosols, whilst level of sedation had little effect.^15^ Sagami *et al* showed that aerosols increased significantly in a plastic enclosure around patients’ heads during UGI endoscopy compared with a control group.^16^ We emphasize that particle counting approaches alone do not directly test whether there is viable virus material in droplets and aerosols – for this, an air sampling approach is required.^1^ However, aerosol and droplet transmission is widely agreed to be the most important physical transmission route for SARS-CoV-2 and so serves as an important proxy indicator for potential infectivity^2^.

Our study aims to characterise aerosol and droplet generation in gastrointestinal endoscopy performed via the mouth, nose or rectum, by quantifying particles across whole procedures and specific events during procedures, and analysing associated risk variables. This information is important in ensuring the safety of GI endoscopy for patients and healthcare workers for both current and future respiratory and gastrointestinal pathogens.

## MATERIALS AND METHODS

### Study design and participants

This is a prospective observational study. Health Research Authority and ethical approval was granted by the Wales Research Ethics Committee prior to the start of the study. We included patients undergoing routine upper and lower GI endoscopy on the lists of thirteen different participating endoscopists at the Endoscopy unit of the Nottingham University Hospitals NHS Trust Treatment Centre between October 2020-March 2021. The inclusion criteria were adult patients >18 years with capacity to consent. For reasons of practicality, whole lists were selected for recruitment and all those on each list who met the inclusion criteria invited to participate. Procedures were performed as they normally would be in clinical practice. Patients chose whether they wanted sedation and endoscopists chose whether they wanted to use carbon dioxide only or water-immersion for insertion during LGI procedures. All UGI procedures were performed with CO_2_ or air for insufflation and intermittent suctioning was used for all per-oral gastroscopies (POG).

Using data from previous studies that measured particle counts and sizes for coughs and sneezes, we calculated that we would need to be able to measure an effect size (Cohen’s *d*) of 1.98 or less to be able to differentiate between relevant events (e.g. cough vs. sneeze). This was computed by comparing the differences in mean particle counts divided by standard deviation for datasets of coughs and sneezes^17-18^. We then computed that measuring this effect size would require at least 5 repeats of each procedure type tested.

To standardise the procedures, we used endoscopy rooms within the same endoscopy suite, which all had room ventilation set at 15-17 air changes per hour, and a similar size, air temperature and humidity levels. We minimized unnecessary airflow for example by not allowing the room doors to be opened during the procedures and only allowing one additional person (the research nurse) in the room. All present in the room wore enhanced PPE which minimised the contribution of additional human aerosol sources.

### Patient and public involvement

The Nottingham University Hospitals’ NHS Trust hosts an NIHR Gastrointestinal & Liver Biomedical Research Centre, through which a Patient Advisory Group has been formed. Three members of this group were recruited to approve the significance of the study and acceptability of the methodology. They also ensured the Patient Information Sheet and Consent form were easily understandable.

### Measurement methodology

We used two pieces of equipment to measure particle sizes. The first was a TSI AeroTrak Portable Particle Counter (model 9500-01) which previous studies have used for respirable particle sizing in medical contexts^19^. This measured particles in six diameter ranges (0.5-0.7μm, 0.7-1.0μm, 1.0-3.0μm, 3.0-5.0μm, 5.0-10.0μm, 10.0-25μm) at a flow rate of 100L/min. A 2m tube (manufacturer provided) was connected to an isokinetic inlet head placed 10cm from the mouth for UGI procedures and approximately 20cm from the anus in LGI procedures using an articulating arm. These distances were chosen for compatibility with previously published studies^13^ and represent an acceptable trade-off between practicality (include access of scope, need to change position of patient) and maximizing aerosol capture (which is known to reduce significantly by 2m in a room with high background particles^21^). The operator’s hands are kept >50cm from the air inlet head to avoid interference from leakage through the endoscope’s suction and air/water control buttons. The effect of the tube length on larger particles is accounted for by a calibration experiment in a room at equilibrium using a 0.02m tube. The second instrument used was an Oxford Lasers VisiSize N60 Spray Characterisation Tool which was used in four POGs. Spray characterisers have previously been used for characterising coughs and sneezes.^17^ The configuration we used allows sizing of particles from ∼10μm to 3.5mm diameter. It is important to consider these two size ranges because respiratory aerosols are thought to be polydisperse, with two size peaks at around 1μm and also around 100μm diameter^21^. The instrument images particles that pass through a small volume located between a laser head and a camera (dimensions 12.6×7.2×50 mm = 4536mm^3^). The instrument is placed such that this volume is located 10cm from the mouth of the patient (see diagram on Patient Information sheet in Supplementary Information, p.8).

During the procedure, an observation camera with a timestamp feature is used to record audio and video for synchronisation purposes. For each procedure, an experienced research nurse recorded information on a case report form containing demographics (age, sex, BMI) and variables determined during the procedure (sedation type, degree of discomfort, use of CO2 or water for LGI procedures, subjective 3-tier estimate of anal tone taken during the pre-procedure digital rectal exam and representing pressure required for insertion, and presence of hiatus hernia). During the procedure, the times of relevant events, beginning when the patient has entered and ending after they have left, are recorded along with the time in seconds. A template is given in the Supplementary Information. Periods of time when there are no significant events, e.g. lengthy examinations without patient movement, are identified and marked as ‘null reference’ events.

Our measurements did not detect statistically significant particle production from rectal insufflation events or injection of water through the scope, which indicates leakage is not likely a significant source of interference. However, our use of intermittent suctioning as per standard protocol may reduce measured particle counts by up to 50%^15^, although we note that those particles not captured by suctioning pose the main airborne viral transmission risk so our measurements have high relevance for infection control.

## Data processing and statistical analysis

### Analysis of full procedure data

We first consider the total particle count across each procedure for 2 particle diameter ranges: 0.5-5μm (aerosols) and 5-25μm (droplets) with patient position changes suppressed. The time period considered starts from either anaesthetic spray (UGI) or intubation (LGI) and ends at extubation. This is compared to a reference window before the procedure starts and is normalised to account for different durations. The fallow period of 20 minutes between procedures should minimise interference from residual particles, but for comparison we also consider an alternative method that uses a background removal technique based on smoothing to estimate particle counts (see Supplementary Information).

### Causal event-based model

We next apply our causal event-based model, which essentially takes a difference in particle counts before and after an annotated event (e.g. cough) to estimate the number of particles produced by the event. For each annotated event, we first estimate the room background particle count immediately before the event by smoothing the data over a 105 second window. We then subtract this background from the raw particle count immediately after the event, averaged over a window of 15-30s. To validate this approach we also apply it to several periods when there was no annotated event and so the difference is expected to be approximately zero.

### Statistical analysis

Building on existing models of aerosol production in the respiratory tract^20^ we use a log-normal distribution to model the distribution of total particle counts across different instances of each event. For the whole procedure data, a t-test is applied to compute p-values. For the causal event model the data distribution can be modelled as the sum of a log-normal and normal distribution to account for negative values of particle counts that can arise from the subtraction step. A Monte-Carlo sampling (or bootstrapping) method is therefore used to provide numerical estimates of p-value^22^.

## RESULTS

### Demographics

Overall, we recorded 48 UGI procedures (37 per-oral, 11 trans-nasal) and 48 LGI procedures (37 colonoscopies, 11 flexible sigmoidoscopies). Due to the recruitment of whole lists, 46% of procedures are consecutive but we do not find any statistically significant correlation between consecutive procedures in terms of particle counts. Of the UGI procedures, 17 performed a volitional cough, 12 performed deep breathing and speaking for reference. Patient variables were as follows. Sex: 52 male, 44 female. Age: range 23-93, median 62 years. BMI: range 16.3-56, median 25.5. Sedation: UGI: 15 midazolam ± fentanyl, 33 unsedated, all procedures used xylocaine throat or nasal spray, LGI: 19 midazolam± fentanyl, 24 Entonox, 4 no sedation. Anal tone: 5 low, 21 medium, 14 high, 8 not recorded. LGI use of CO_2_ vs water: 42 CO_2_, 6 water. Discomfort: 56 low, 33 medium, 3 high, 4 not recorded. UGI hiatus hernia: yes 12, no 36. Smoker: 14 yes, 82 no. LGI diverticular disease: 40 none, 4 mild, 4 extensive.

### Whole procedure analysis

Over the full range of particle sizes (0.5-25μm) and normalised to procedure duration, POG produced significantly higher particle counts than the reference background (1.96x, 95%CI:1.61-2.38, *p*<0.001, n=37) as did TNE (2.00x, 95%CI:1.53-2.62, *p*<0.001, n=11). However, directly comparing POG and TNE, we find that POG produces significantly more particles (1.99x, 95%CI:1.28-3.12, p<0.01). LGI procedures (with patient position changes excluded) were significantly higher than the reference background (1.34x, 95%CI:1.14-1.59, *p*<0.001, n=48), but less so than UGI procedures. When applying background removal, we find that the ratios compared to the reference window increase approximately by a factor of 3 with the main trend preserved. However, we find that the ratio between POG and TNE becomes non-significant (p=0.411), which indicates the importance of slow but continuous production of aerosols during POG, as opposed to event-driven ‘spikes’, that are removed by our background subtraction technique. If we exclude the anaesthetic spray from our analysis we find the ratios compared to reference window are reduced by about 36% for both POG and TNE but are still significant (p<0.01, p<0.05 respectively), indicating the importance of anaesthetic spray as a driver of particle production, which agrees with our event-based analysis.

The absolute number of particles is on average less for POG than for LGI procedures (0.71×10^8^ vs 1.69×10^8^), but is greater when procedure duration (intubation to extubation) is taken into account: POG produce particles at a rate of 13.9×10^6^ (95% CI:7.3×10^6^-20.5×10^6^) per minute/m^3^ vs 8.8×10^6^ (95%CI:4.0×10^6^-13.6×10^6^) per minute/m^3^ for LGI procedures excluding position changes. The median duration of recorded procedures is 7.2 mins for UGI and 24.7 mins for LGI. Within the LGI procedures we find no significant difference between colonoscopy and sigmoidoscopy in terms of particle production rate (p=0.168) although the absolute number of particles is larger for colonoscopy (1.86×10^8^ vs 1.23×10^8^) because the procedures are longer (median 26.0 minutes vs 10.3 minutes)

For particles >5μm in diameter we find that LGI procedures are no longer significant relative to the background (*p*=0.082). For particles <5μm in diameter we find all procedure types are significantly higher than reference background (POG: 1.99x, TNE: 2.09x, LGI: 1.34x, *p*<0.001). The particle counts, normalised to procedure duration, relative to the reference background are summarised in Figure 1.

**Figure 1:**
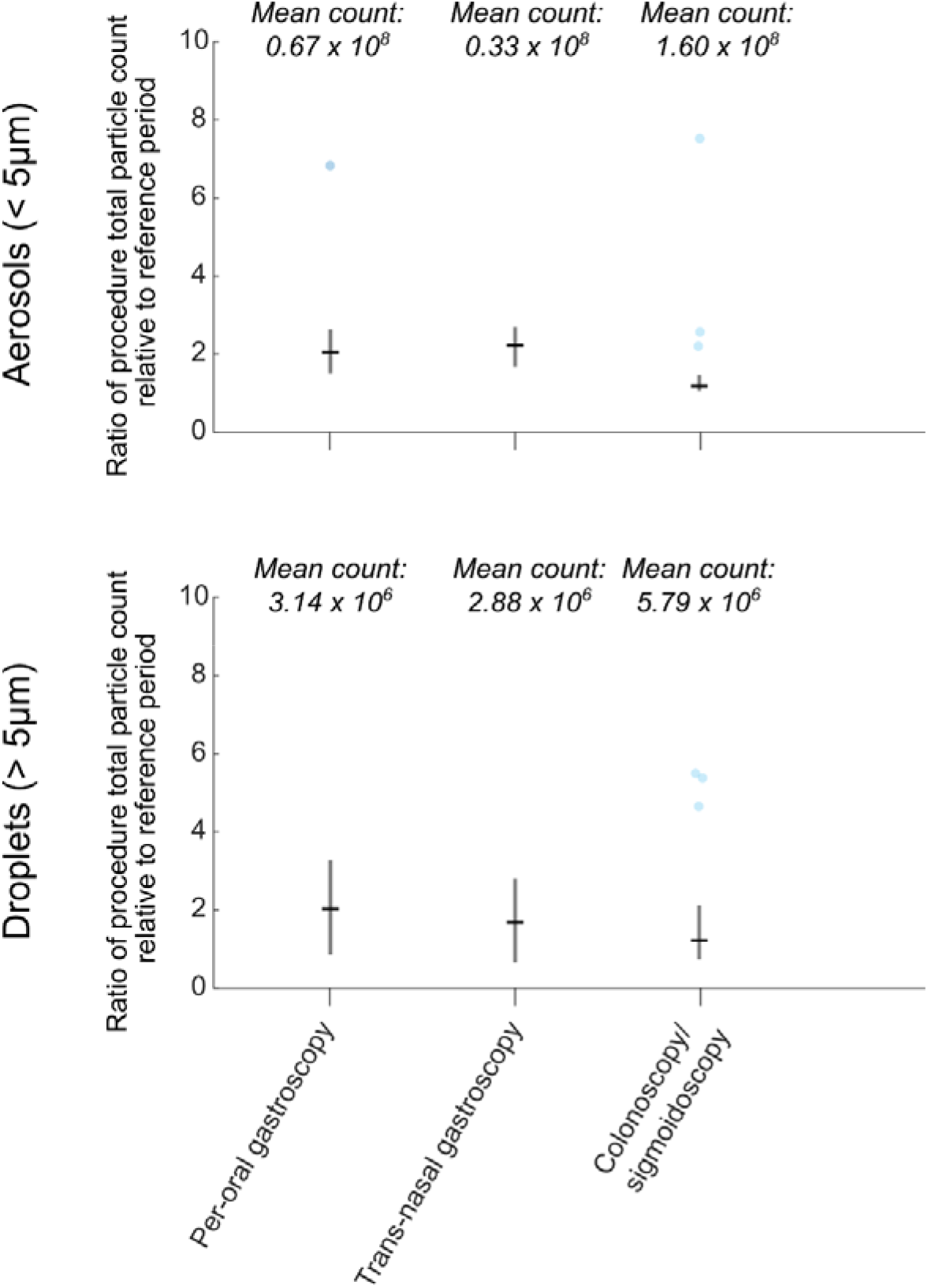
Ratios of particle counts over whole procedures relative to a reference period before the start of the procedure (normalised to procedure duration). White circles indicate median values. Raw mean counts (not normalised to procedure duration) are shown above.

Regarding variables, the only significant result for LGI procedures was patient discomfort rated ‘high’ resulted in more particles than discomfort rated ‘low’ (6.3x, 95%CI:1.6-25.3, *p*<0.01). For UGI procedures there was a small statistically significant (p < 0.05) negative correlation between particle count and age (r^2^ = 0.09). Other variables, including BMI, use of sedation, use of CO_2_ or water for insertion, were not found to have significant effect.

### Causal event-based analysis

We next consider individual events, shown in Fig.2. For UGI procedures we find the following events significant relative to the room background particle count: nasal intubation (10.9x, 95%CI:0.68-256.6, *p*<0.05, n=11), oral extubation (37.5x, 95%CI:6.3-619.3, *p*<0.001, n=35), nasal extubation (32.0x, 95%CI:4.0-612.4, *p*<0.01, n=11), coughing/gagging during oral endoscopy (25.8x, 95%CI:3.5-483.5, *p*<0.01, n=28), coughing/gagging during TNE (20.0x, 95%CI:2.3-398.0, *p*<0.01, n=17), forced coughing (7.5x, 95%CI:0.67-143.7, *p*<0.05, n=17), deep breathing (15.7x, 95%CI:1.4-329.8, *p*<0.05, n=12), anaesthetic nasal spray (40.1x, 95%CI:4.6-737.1, *p*<0.001,n=13), anaesthetic throat spray (150.0x, 95%CI:19.4-2697.0, *p*<0.001, n=30). Oral intubation is not significant (*p*=0.443, n=32) nor is speaking at low volume (*p*=0.170, n=12), which is consistent with previous studies.^6^

**Figure 2:**
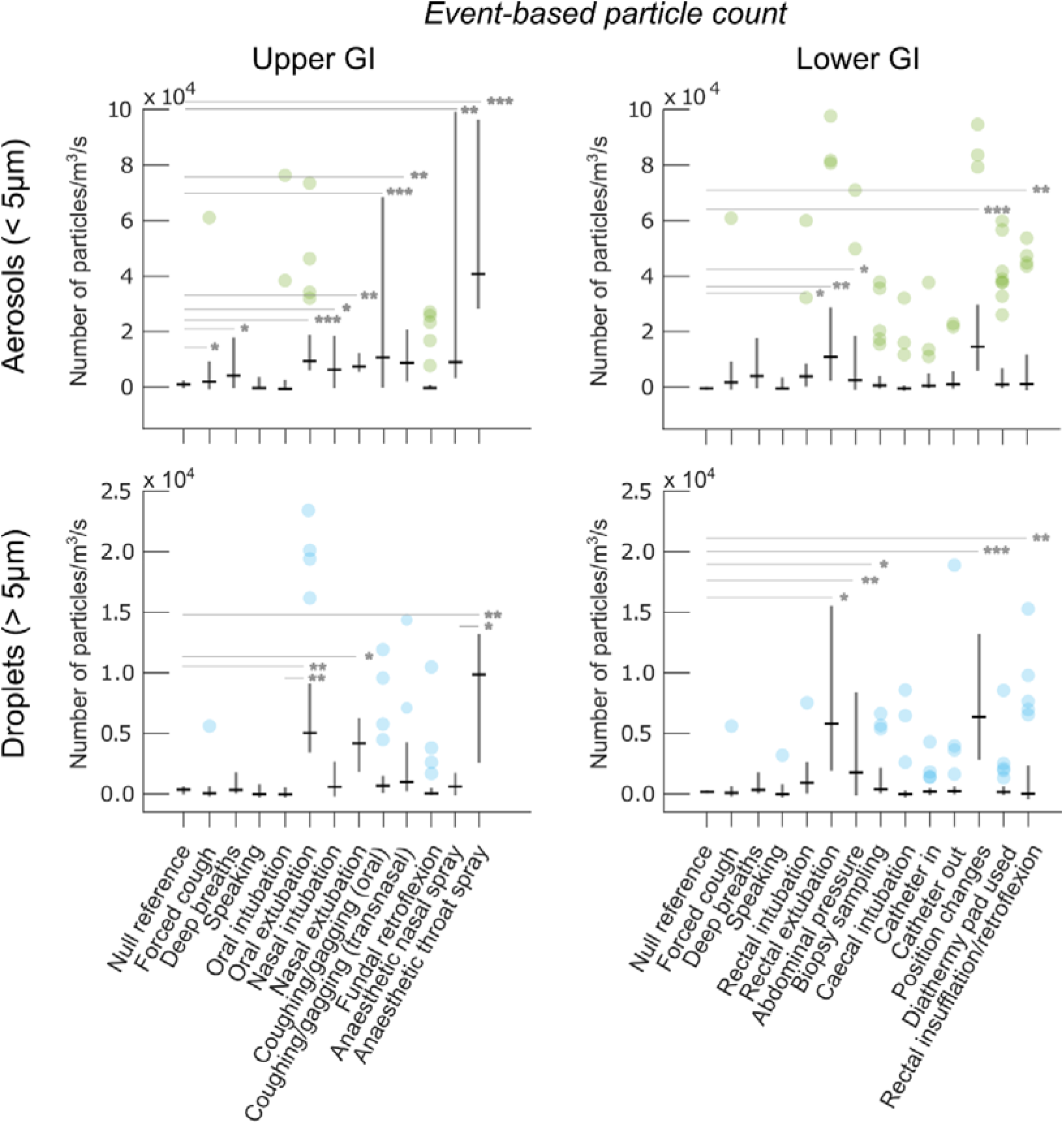
Particle production by individual events measured during upper and lower GI procedures. Numbers of recorded events are given above. Black dashes represent medians. * = p<0.05, ** p<0.01, *** p<0.001. For readability, only a selection of salient statistical relationships are shown.

For LGI procedures we find several events that are significant relative to the room background particle count: rectal intubation (9.9x, 95% CI:1.5-112.1, *p*<0.05, n=45), rectal extubation (27.2x, 95% CI: 6.0-317.7, *p*<0.01, n=49), application of abdominal pressure (9.6x, 95% CI:1.3-130.1, *p*<0.05, n=22), patient position changes (34.9x, 95% CI:9.5-382.8, *p*<0.001, n=98) and rectal insufflation/retroflexion (7.7x, 95% CI: 1.5-98.8, *p*<0.01, n=39). We observe that rectal extubation produces significantly more particles than intubation (3.3x, 95% CI: 1.1-8.6, *p*<0.05). Biopsy sampling, insertion/removal of catheters, water injection and the use of diathermy cutting are not significant.

### Comparison to volitional coughing

To examine the relevance for potential airborne pathogen spread, we next compare the events to volitional coughing, in line with previous work.^6^ For UGI procedures, we find the following events statistically indistinguishable from the mean volitional cough: coughing/gagging during transnasal (p=0.116), deep breaths (*p*=0.187), speaking (*p*=0.230). However, some events produce significantly more particles: oral extubation (4.0x, 95%CI: 1.6-27.1, *p*<0.01) anaesthetic throat spray (10.5x, 95%CI: 1.2-80.1, *p*<0.05), nasal extubation (4.2x, 95%CI: 0.9-20.5, *p*<0.05), coughing/gagging during oral (3.5x, 95% CI: 0.8-20.5, *p*<0.05), anaesthetic nasal spray (5.5x, 95%CI: 0.9-43.9, *p*<0.05).

For LGI procedures particle generation is comparable to a forced cough for intubation (*p*=0.404), extubation (*p*=0.171) and abdominal pressure (*p*=0.212), but significantly more particles are produced for rectal extubation (5.9x, 95%CI: 1.5-40.4, *p*<0.01) and position changes (7.0x, 95% CI:2.0-40.9, *p*<0.05).

### Particle size analysis

The size range of particles associated with each event is shown in Fig.3. For UGI procedures, oral extubation produces particle sizes significantly larger than volitional coughing (2.2μm vs 0.32μm, *p*<0.001 for both), whilst particle sizes are similar for involuntary coughing/gagging (oral: 0.44μm, *p*=0.36, nasal: 0.68μm, *p*=0.35). Both anaesthetic throat spray (*p*=0.09) and nasal spray (*p*=0.31) produce particles statistically similar in size to coughing. For LGI procedures, rectal extubation produces particles of a similar mean size to oral extubation (2.0μm, *p*=0.226). Position changes of the patient produces particles comparable to rectal extubation (1.7μm, *p*=0.30).

**Figure 3:**
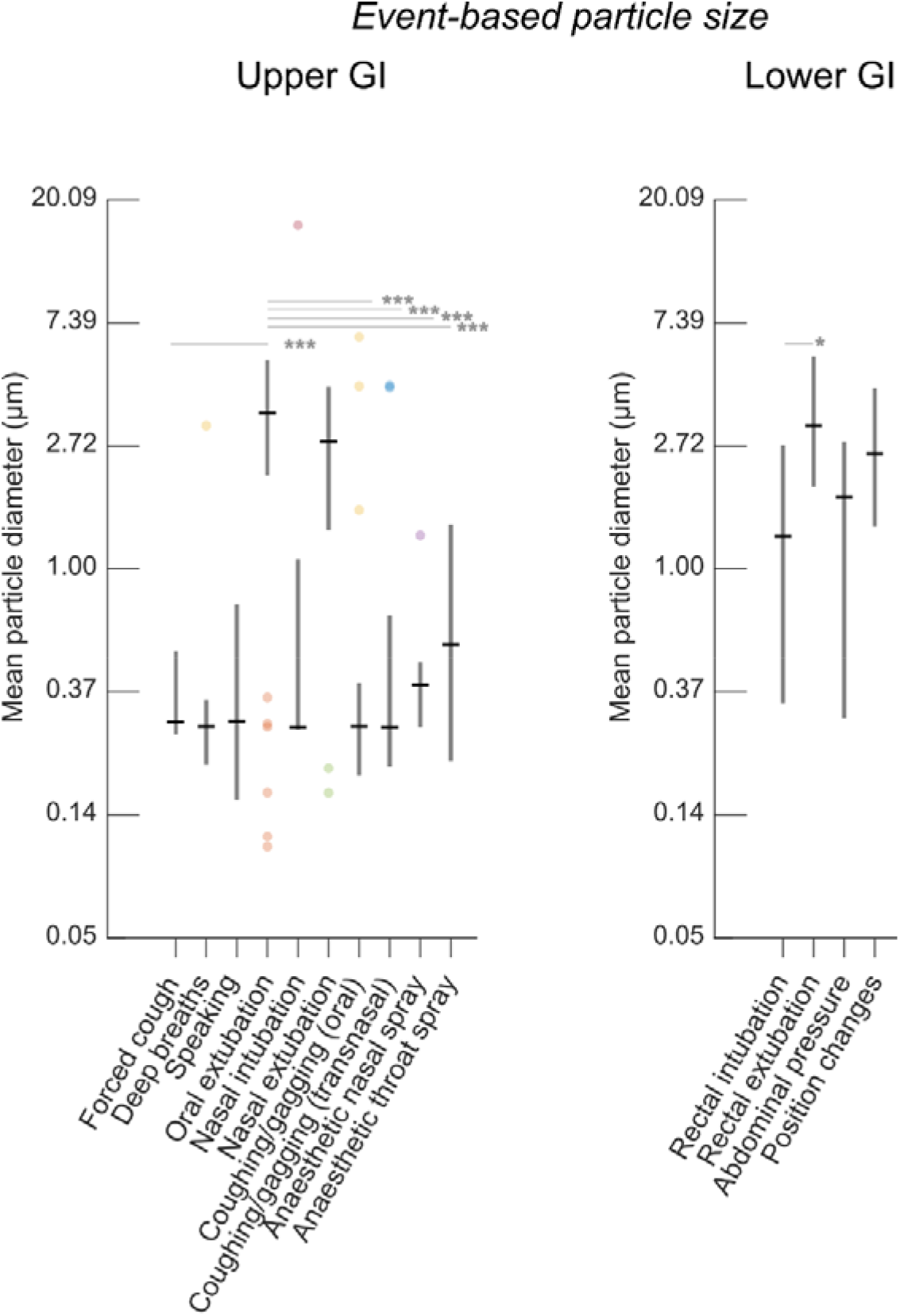
Particle size distribution for statistically significant particle generating events. * = p<0.05, ** p<0.01, *** p<0.001. Note that for readability, only a small selection of salient statistical relationships are shown.

To examine the effect of larger particles (>10μm), we used a spray characteriser to record four separate POGs. We observe a strong temporal correlation in particle counts between this instrument and the AeroTrak which confirms that both instruments are recording the same particle producing events (see Supplementary Figure 1). For the cases examined there is insufficient data for full statistical analysis but we found that oral extubation and fundal retroflexion produce particles up to 300μm (mean measured diameter 32μm), whilst coughing/gagging does not produce detectable particles in this range (see Supplementary Figure 1).

### Impact of variables

Finally, we analyse the effect of measured variables on event-based particle production. In the presence of a hiatus hernia, there was a much larger increase in particle generation during volitional coughing (23.2x, 95%CI: 1.6-346.7, *p*<0.05), which may warrant further investigation. We also note that on average there are 2.5x as many coughing/gagging events per procedure for patients who have a hiatus hernia (*p*<0.05). For UGI, there is limited impact of variables on particle size.

For LGI procedures, the variables have minimal effect on rectal intubation and rectal extubation: sedation, anal tone and age are not statistically significant.

## DISCUSSION

This is the first study to report that both TNE and LGI endoscopy are aerosol and droplet generating. We are also the first to report on defined particle generating events and associated particle sizes within procedures performed via the mouth, nose and rectum. Both POG and TNE should therefore be classed as AGPs, whilst the classification of LGI endoscopy depends on the definition of AGP used. For UGI endoscopy, in some countries the use of patient facemasks is becoming widespread though there have been few studies quantifying their impact on aerosol and droplet generation^23^. A randomised study recently found that a mouthpiece did not reduce smaller size particles (<1.0μm), which in the context of our findings may suggest limited reduction of risk^24^: further studies in this area are certainly warranted. However, our aim here is to establish what can be expected during standard endoscopy, without the use of facemasks, that is still the usual practice in most endoscopy units worldwide.

With regards to POG, our results confirm those of previous studies showing this is an AGP, producing particles at double the background level. The most significant contributing event is local anaesthetic throat spray application, which generates ten times the number of particles compared to a volitional cough, with an average particle size in the aerosol range. By comparison, a recent study showed that controlled endotracheal intubation and extubation in asymptomatic patients generate only a fraction of the aerosols generated by volitional coughing.^25^ The particles recorded with throat spray application are potentially infectious, as they would have rebounded from the patient’s oropharynx or occasionally, been contributed by coughing induced by the throat spray. There is additional risk because the throat spray is applied face-on with the patient. It is therefore important that barrier methods such as face shields or goggles are used whilst applying throat and nasal spray.

Extubation is the second most particle generating event in POG and is also significantly more particle generating than volitional cough. However, a higher proportion of particles is in the droplet range (and reaches up to 3000μm), which has a lower risk for airborne transmission. This is understandable as both insufflation in the oesophagus and the movement of the wet shaft of the endoscope on extubation will generate particles.^8^ Coughing/gagging are also significant generators of particles, and is predictably comparable to the level of particles produced by volitional coughing, although we did not find that the use of sedation reduced particle counts over the whole procedure. The usefulness of suctioning, described by Chan *et al*,^15^ cannot be commented on in our study, as intermittent suctioning was applied in all of our cases.

Interestingly, during volitional coughing, we found the presence of a hiatus hernia gives increased levels of particles, with an average size in the aerosol range. This may be due to the loss of the physiological lower oesophageal sphincter, which would enable aerosols to be expelled unimpeded from the stomach and out of the mouth as abdominal muscles are contracted during coughing. Previous studies have found negligible impact of sedation on aerosol production, in agreement with our findings, but have found significant positive correlation with BMI which we did not observe^15,16^. We expect that this is because of increased inter-patient variation in our study caused by looser confinement of aerosols – for example, one previous study placed the patient’s head in an enclosure. However, we purposefully designed our study with looser constraints to enable direct comparison between UGI and LGI (for which building a suitable enclosure would be challenging) and to replicate realistic procedure conditions so as to measure aerosol exposure likely to be experience by healthcare workers.

TNE has been suggested by some as a non-AGP method for performing UGI endoscopy, although the generation of aerosols from intranasal application of spray has already been suggested.^26-27^ Our results show that TNE is an AGP and produces particles predominantly in the aerosol range, which may also have implications for similar otolaryngology procedures. Nasal spray application, nasal intubation and nasal extubation were all associated with significant spikes of particles. TNE generates approximately half the level of particles than POG; therefore, if used with additional mitigating strategies (avoidance of nasal spray, barrier methods) TNE could potentially become a non-AGP procedure.

With regards to LGI endoscopy, our study shows the absolute levels of particles produced are greater than UGI procedures, but are about one third lower when taken per unit time. Although there would be a greater exposure to aerosols in LGI procedures due to longer procedures, these are therefore more likely to be cleared in well ventilated rooms. We recognise that COVID-19 is primarily a respiratory pathogen, and feco-oral transmission has not been proved. The risk from LGI procedures is likely to be considerably lower than equivalent aerosols generated by UGI procedures. However, it should be noted that infection of intestinal cells and viral replication has been shown,^27^ and SARS-Cov-2 RNA has been detected in stools,^28^ whilst there are also implications for other types of gastrointestinal pathogens. There have been attempts to mitigate aerosol and droplet diffusion during colonoscopy using specially designed shorts with a diaphragm to pass the colonoscope.^29^

An important source of interference to consider for LGI endoscopy occurs during patient position changes. We observe that turning a patient in the bed before the procedure has even started results in a large spike in measured particles, which is probably due to air movement and the rubbing of materials. The clinical relevance of position changes is therefore difficult to interpret but we are able to identify, isolate and exclude these from analysis where appropriate.

In this study, we have characterised aerosol and droplet generation from the different routes of GI endoscopy. We emphasise, however, that aerosols may not necessarily contain viable virus material and so their generation does not equate to infectivity of the procedures themselves. This depends on multiple factors, including which part of the patient the particles are being generated from; particles from the oral and nasal cavities are likely to have a much higher potential infectivity risk compared to those from the large bowel. As the infectivity of procedures is not established, we therefore suggest adequate PPE (including high-efficiency masks) and sanitization of floors and surfaces (to prevent resuspension of aerosols) is used for all GI endoscopy where there is a high population prevalence of COVID-19.

## CONCLUSION

Our study shows endoscopic procedures performed via the mouth, nose or rectum all generate aerosols and droplets, and that individual events produce greater or comparable levels of particles compared to volitional cough. For UGI endoscopy, our results suggest aerosol generation can be greatly reduced by avoiding or finding alternatives to throat spray, and by performing TNE, but TNE is still an AGP and further mitigating strategies should be applied. LGI endoscopy produces more particles per procedure, but is less particle generating per unit time and produces more particles in the droplet range. The main contributing events are rectal extubation, application of abdominal pressure and rectal intubation. More studies are needed to evaluate mitigation strategies and to characterise the infectivity of these procedures themselves.

## Data Availability

Data associated with this publication is available at http://dx.doi.org/10.17639/nott.7112  Code used for data analysis in this publication can be found at https://github.com/gsdgordon/aerosols

## ACKNOWLEDGEMENTS

The authors thank Guru Aithal for critically reviewing the manuscript; Martin James and Bu Hayee for reviewing the study protocol; Matthew Sanderson, Andy Wragg, Nottingham University Hospitals Research and Innovation, University of Nottingham Biomedical Research Centre, Karren Staniforth, Laura Leman, Nina Duffy, Allison Ball and the Endoscopy Unit Staff in their support of the development of this study; the NIHR Aerosol Generating Procedures Group for their support during the study; Tina Rodriguez, Paul Brocklebank, Mirela Pana, Sabina Beg, Stefano Sansone, James Catton, Emilie Wilkes, Lorraine Clark, Andrew Horton, John White, Suresh Vasan Venkatachalapathy, Aida Jawhari, Ioannis Varmpompitis, and Muthuram Rajaram for performing endoscopic procedures in this study; and Olympus for loan of the trans-nasal endoscopes

## PATIENT CONSENT

Obtained

## ETHICS APPROVAL

Wales Research Ethics Committee

